# Safety and Effectiveness of SA58 Nasal Spray against SARS-CoV-2 family transmission: an exploratory single-arm trial

**DOI:** 10.1101/2023.03.19.23287462

**Authors:** Lianhao Wang, Rui Song, Yuansheng Hu, Gang Zeng, Keqiang Sun, Jianfeng Wang, Yafeng Bao, Yun’ao Zhou, Long Cheng, Can Wu, Junfan Pu, Xing Han, Junlan Wu, Ronghua Jin, Qiang Gao

## Abstract

**Background:** This study has assessed the protective effect of a new Anti-COVID-19 SA58 Nasal Spray (SA58 Nasal Spray) against SARS-CoV-2 infection under continuous exposure.

**Methods:** This is an exploratory open-label, single-arm trial. To evaluate the safety and effectiveness of SA58 against SARS-CoV-2 family transmission, SA58 was administered to all enrolled family contacts at 3∼6-hour intervals. The frequency of administration and adverse events (AEs) were self-reported by online questionnaire, and RT-PCR tests were used to diagnose SARS-CoV-2 infection. The effectiveness was assessed in comparison to a contemporaneous control group whose information was collected through three follow-up visits. Total effectiveness and single-day effectiveness were calculated.

**Results:** The incidence of SARS-CoV-2 infection was 62.9% (44/70) in the experimental group and 94.8% (343/362) in the control group. Using SA58 nasal spray at least three times per day could possibly reduce the risk of household transmission of SARS-CoV-2 by 46.7%∼56.5%. The incidence of AEs was 41.4% and the severity of all AEs was mild.

**Conclusion:** Even under the scenario of continuous exposure to SARS-CoV-2, SA58 nasal spray remained effective in blocking viral transmission and was well tolerated.

## BACKGROUND

The past three years saw the development of various types of vaccines against COVID-19 at an unprecedented pace, with 13 vaccines having been added to the WHO emergency use list[1]. However, efficacy of all types of COVID-19 vaccines is progressively declining with the constant variation of SARS-CoV-2, especially efficacy against infection[2, 3]. Worldwide, the Omicron variant pandemic has further confirmed its greatly increased ability of transmissibility and immune escape[4]. A cohort study from England showed that the risk of severe outcomes is substantially lower for Omicron than for Delta and a significant variation with age[5]. Surveillance data from Hongkong also indicated that Omicron was associated with a lower rate of severe symptoms and mortality in healthy adults and adolescents who had received 2 or more doses of vaccine, but these rates remained high in the elderly and in people with underlying medical conditions[6]. Therefore, we still need to further explore ways to prevent COVID-19 infection, especially with a broad-spectrum vaccine or a drug that could protect against as many variants as possible.

Neutralizing antibodies, which could effectively block virus entry into host cells, are urgently needed for intervention against COVID-19[7]. However, numerous approved neutralizing antibodies are designed to target the S protein structure of SARS-CoV-2 in binding cells[8]. Thus, the effectiveness of neutralizing antibodies is limited by the sensitivity of the predominant circulating strains to these neutralizing antibodies due to their rapid mutation[9]. With the worldwide transmission of Omicron, all neutralizing antibodies used for the treatment of COVID-19 were virtually ineffective, especially when patients were infected by subvariants of BQ and XBB[10]. Therefore, the development of broad-spectrum neutralizing antibodies which could effectively neutralize current and future major variants, has become one of the potentially alternative strategies to reduce mortality of COVID-19.

SA58 Nasal Spray, a broad-spectrum anti-COVID-19 candidate mAb, is developed by Sinovac Life Sciences Co., Ltd. It has been shown to potently neutralize ACE2-utilizing sarbecoviruses, including circulating Omicron subvariants in in vitro neutralizing and in animal challenge studies[11]. Two previous IITs (investigator-initiated trials) have indicated that the effectiveness against SARS-CoV-2 infection of SA58 nasal spray was 77.7% (95% CI: 52.2% - 89.6%) and 61.83% (95%CI 37.50%-76.69%) in people with non-continuous exposure to COVID-19 separately, and the safety results indicated that the SA58 Nasal Spray was well tolerated[12, 13]. The government of China announced to adjust the dynamic zero-Covid policy in December 2022, which means many Chinese people would get SARS-CoV-2 in a very short period. The implementation of home quarantine policy has also led to a greatly increased risk of household transmission. Considering that continuous exposure to COVID-19 is one of the high-risk settings for transmission of SARS-CoV-2, we recruited families within Sinovac staffs and their relatives or friends who had at least one COVID-19 case in their families and in whom all members were willing to use SA58 nasal spray. Considering that Sinovac employees are vulnerable subjects in this research, only those who spontaneously contact with the Sinovac’s clinical R&D center could be recruited. The personal information of subjects is also strictly confidential to ensure that it will not be disclosed. In order to figure out the characteristics of SARS-CoV-2 in a setting of household transmission, we also conducted three telephone follow-up visits for SARS-CoV-2 infected households without using SA58 nasal spray as an external control group. The aim of this study is to evaluate the effectiveness of SA58 nasal spray against SARS-CoV-2 infection in the scenario of continuous (intra-family) exposure to COVID-19.

## METHODS

### Study design

This exploratory, open-labeled, single-arm study evaluated the effectiveness and safety of the SA58 nasal spray in household transmission in Beijing, China. In the experimental group, we recruited 62 families among Sinovac’s employees and their friends or relatives from Nov 9^th^ to Nov 24^th^, 2022. In the control group, we recruited 154 families whose member had already been infected with SARS-CoV-2 (confirmed by RT-PCR or antigen test) and had not used SA58 nasal spray. We conducted three telephone follow-up visits on Nov 13^th^, Nov 17^th^ and Nov 22^nd^, 2022, respectively, for these families to figure out the characteristics of household transmission. All participants in this study were voluntary and fully informed consent before enrollment. The clinical trial protocol and informed consent form were approved by the Ethics Committee of Beijing Ditan Hospital, Capital Medical University (Reference No. DTEC-YW2022-024-01). The study was registered with ClinicalTrials.gov (NCT05667714).

### Experimental group

Eligible participants were healthy adults who were willing to sign informed consent with legal identification. The inclusion criteria included (1) at least one family member had a positive result by RT-PCR or antigen test of SARS-CoV-2 within the previous 72 hours from Nov 9^th^ to Nov 25^th^, 2022. (2) all members were willing to participate in this study and use the SA58 nasal spray. The exclusion criteria included 1) individuals with known history of severe allergies or reaction to any component of inhaled SA58 nasal spray. (2) those currently pregnant, lactating, or expected to be pregnant during the study period. (3) those who participated in any kind of clinical trials of SARS-CoV-2 neutralizing antibody injections in the preceding 180 days before screening. (5) were unable to take nasal spray inhalation. (6) had severe neurological disease (e.g., epilepsy, convulsions, or seizures) or psychosis, or family history of psychosis. (7) had any other significant chronic disease, disorder, or finding that, in the judgment of the investigator, significantly increased the risk to the participant because of participation in the study, affected the ability of the participant to participate in the study, or impaired interpretation of the study data.

We provided RT-PCR sampling tubes and SA58 nasal spray to subjects when they were recruited into the experimental group. All family members were asked to self-collect throat swabs at Day 0 (delivery date of SA58), Day 1, Day 2, Day 5, Day 8. In addition, participants were asked to fill in an online questionnaire for the frequency of SA58 use and adverse events once a day. Families that met either of the following two criteria could conclude the study:(1) all members in a family have been confirmed to be infected by SARS-CoV-2 (positive results by RT-PCR or antigen test of SARS-CoV-2) (2) In the families where part of members were infected by SARS-CoV-2, two consecutive RT-PCR tests of infected members were negative, which means the uninfected members were not continuous exposed to SARS-CoV-2. Thus, these family members no longer need to self-collect the throat swabs and finish the questionnaire. A follow-up visit was performed on Jan 3^rd^, 2023, to confirm that the subjects with RT-PCR negative results had not developed any Covid-19 related symptoms throughout the study.

### Control group

We had a preview of COVID-19 infections in Sinovac employees’ families. For families who did not join in the experimental group and were willing to undergo subsequent follow-up, we conducted three telephone follow-up visits on Nov 13^th^, Nov 17^th^ and Nov 22^nd^, 2022, respectively. We mainly collected the number of family members, whether they lived together, the confirmed date of first COVID-19 case (Nov 1^st^ – Nov 13^th^, 2022), number of subsequent cases and whether SA58 nasal spray was used. The families whose member had used SA58 nasal spray before and the subjects who did not live with others were excluded. In the control group, the date of positive RT-PCR or antigen test of the first COVID-19 case in his/her family was set as Day 0. Time intervals were calculated by subtracting the date of telephone follow-up visits from the date of Day 0.

### SA58 nasal spray

The SA58 nasal spray manufactured by Sinovac is a liquid medicine containing 5 mg/ml of antibodies which has been verified to be able to neutralize many variants of SARS-CoV-2 in vitro. Each administration of the drug consisted of two sprays with one spray in each nostril, and a total of 1 mg antibody being administered. Three to six administrations of SA58 were recommended at an interval of 3-6 hours per administration in a day, with the last administration given before going to sleep.

### Effectiveness against SARS-CoV-2 infection assessment

The date of the SA58 nasal spray delivery was taken as Day 0. The RT-PCR results of Day 0 and Day 1 were considered to be the baseline infection status of the subject. Only the subjects with these two negative RT-PCR results were considered baseline uninfected and included in analysis. First, we calculated the incidence of SARS-CoV-2 infection in both experimental group and control group, and the effectiveness of SA58 nasal spray in reducing SARS-CoV-2 infections over the entire observation period.

Second, considering that the effectiveness of SA58 nasal spray is closely related to the regular medication use by the subjects, we intended to assess the single-day effectiveness of SA58 nasal spray against SARS-CoV-2 infection by the number of daily new infections in the experimental group and the control group. Since the dates of RT-PCR in experimental group were Day 0, Day 1, Day 2, Day 5, Day 8…, the detection peak of SARS-CoV-2 infection was on the 2^nd^, 5^th^, and 8^th^ day. Under the assumption that the spread of the disease is relatively uniform over the three-day intervals, we averaged the daily number of confirmed COVID-19 cases in three-day periods. For example, the adjusted number of confirmed COVID-19 cases in Day i was equal to the average value of the number of confirmed COVID-19 cases in Day i-1, Day i and Day i+1. As for the daily infection rate of the control group, we didn’t perform this data processing because it didn’t be influenced by the frequency of RT-PCR. We calculated the single-day effectiveness against SARS-CoV-2 infection to see the change in effectiveness over time and the frequency of SA58 use.

### Safety analysis

We summarized the adverse events (AEs) collected by the questionnaire and excluded the AEs occurring after the positive RT-PCR results of the subjects in an attempt to calculate the incidence of AEs during the uninfected period. In most cases, there was a lag between SARS-CoV-2 infection and RT-PCR detection, we calculated the adjusted incidence of AEs after excluding AEs within two days of the first positive RT-PCR as well, to exclude the AEs were probably related with COVID-19 infection.

### Statistical analysis

The total effectiveness (*TE*) of the SA58 nasal spray was evaluated by calculating the infection rate (IR) of COVID-19 cases. The infection rate of each group (*Cumulative Incidence*_*i*_) was defined as the numbers of COVID-19 infections divided by the number of the group as per formula (1). The total effectiveness against infection (*TE*) was evaluated by the formula (2).

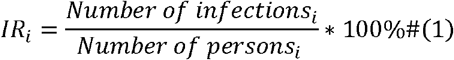

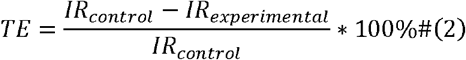

The daily infection rate at Day t, *DIR*_*i,t*_ was calculated as the number of new infections at Day t divided by the number of susceptible people at the start of Day t as formula (3). The single-day effectiveness at Day t (*SE*_*t*_) of SA58 nasal spray was evaluated by the formula (4).

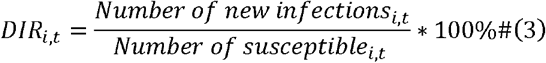

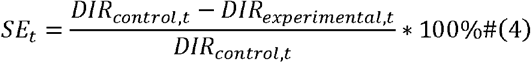

Kaplan–Meier curves were also presented for experimental and control groups, with hazard ratios (HR) calculated by using Cox proportional hazard models. AEs were summarized descriptively as frequencies and percentages by type of event and severity. All statistical analysis was conducted by the SPSS 26.0 and Excel 2022.

## RESULTS

### Study population

From Nov 9^th^ to Nov 24^th^, we recruited a total of 166 subjects in 56 families into the experimental group. After excluding those who did not use SA58 nasal spray (N=4), had no RT-PCR results (N=3), had unknown exposure status (N=5), were the first COVID-19 case in the family (N=44), and were probably already in the incubation period of infection (positive RT-PCR results on Day 0 or Day 1, N=40), 70 subjects in 40 families were included into the analysis set (Figure 1). The maximum exposure period was 13 days. The highest daily average frequency of SA58 use was for Day 1 with 4.1 times per day. The daily average frequency continued to decrease from Day 1 to Day 8, with a minimum of 1.9 times per day (Figure 2).

**Figure 1.**
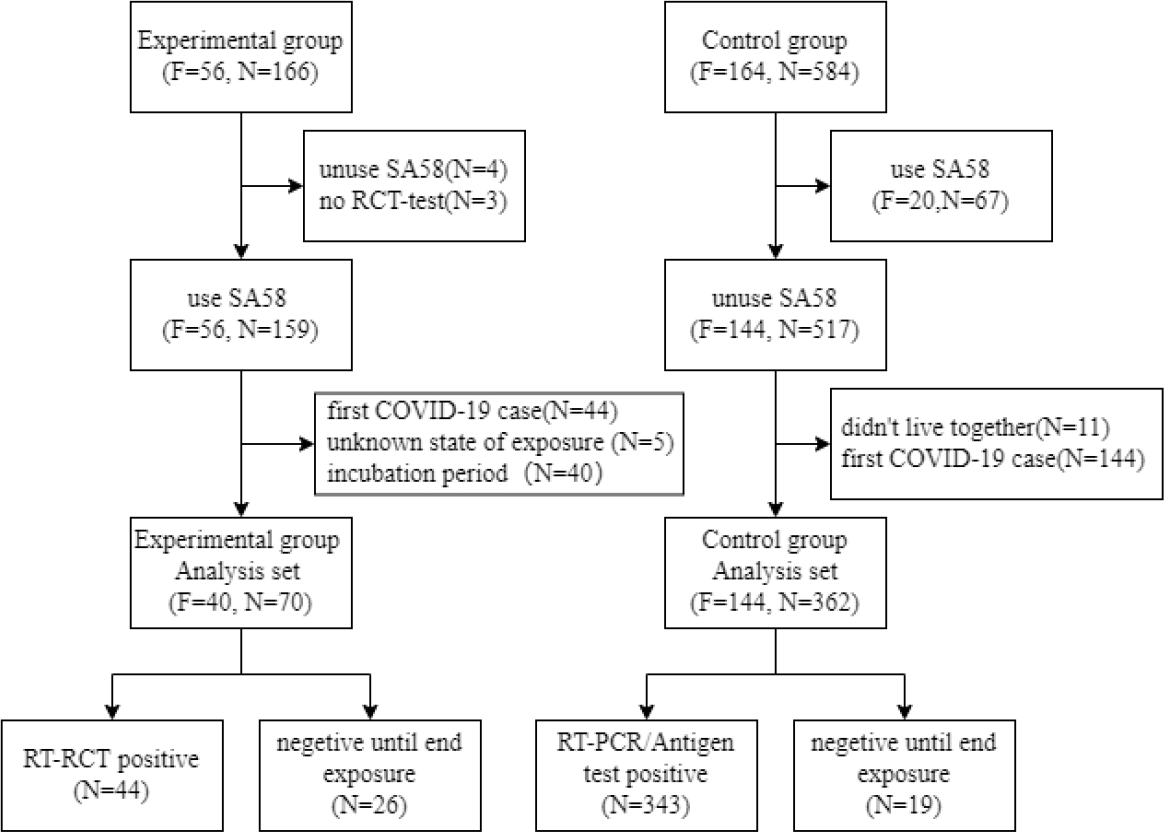
Study design and the number of subjects. F means the number of families and N means the number of people.

**Figure 2.**
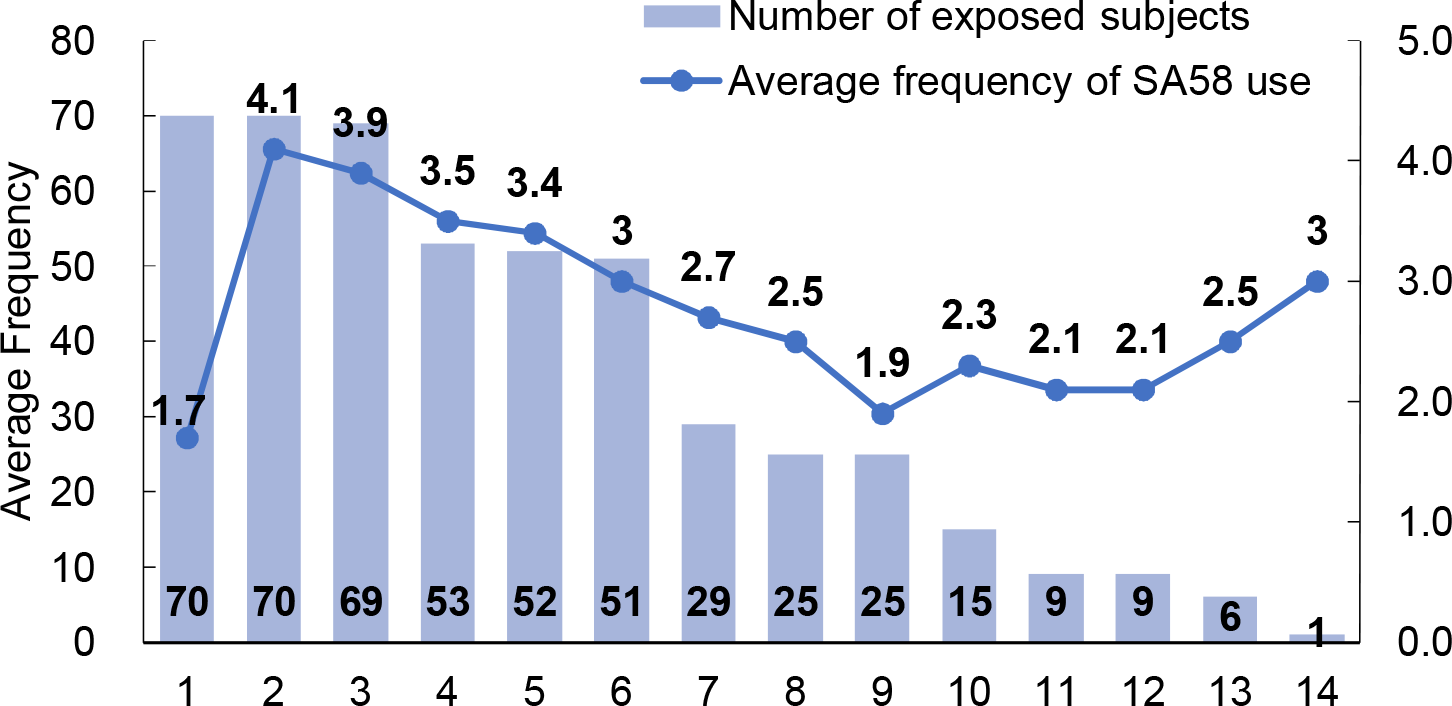
Trends of the number of people with continuous exposure to COVID-19 and the average frequency of SA58 use over time in the experimental group.

For the control group, we recruited a total of 584 subjects in 164 families. We excluded subjects who used SA58 nasal spray before (N=67), did not live with other family members (N=11), and were the first COVID-19 case in the family (N=144) by the information collected at three follow-up visits. Finally, 362 subjects in 144 families were included into the analysis set (Figure 1). The longest period of continuous exposure to COVID-19 was 16 days.

### Effectiveness against SARS-CoV-2 infection

In the experimental group, a total of 44 cases (62.9%) had positive results of SARS-CoV-2 RT-PCR results during the observation period, and the other 26 subjects (37.1%) were verified to be persistently negative when other family members recovered from SARS-CoV-2 infection. The cumulative incidence of infections in the experimental group was 62.9%. In the control group, the infection rate was 94.8% (343/362). The total effectiveness against SARS-CoV-2 infection of SA58 nasal spray was 33.79%.

The single-day effectiveness against infection of SA58 nasal spray was relative stable from Day 2 to Day 5 with the rate of 46.7%∼56.5%. After Day 5, the single-day effectiveness varied greatly and did not show obvious regularity (Table 1).

**Table 1.** Daily infection rate of subjects in experimental group (*DIR*_*exp,t*_) and control group (*DIR*_*con,t*_) and the single-day effectiveness against SARS-CoV-2 infection (*SE*_*t*_).

| Day | $DIR_{exp,t}$ | $DIR_{con,t}$ | $SE_t$ |
| --- | --- | --- | --- |
| 1 | 6.19% | 6.90% | 10.24% |
| 2 | 7.11% | 15.74% | 54.85% |
| 3 | 8.20% | 15.38% | 46.72% |
| 4 | 11.90% | 22.94% | 48.11% |
| 5 | 14.19% | 32.58% | 56.45% |
| 6 | 15.75% | 20.83% | 24.41% |
| 7 | 5.61% | 22.11% | 74.63% |
| 8 | 6.93% | 33.78% | 79.49% |
| 9 | 7.45% | 6.12% | -21.63% |
| 10 | 4.60% | 15.22% | 69.79% |
| 11 | 1.20% | 28.21% | 95.73% |
| 12 | 2.44% | 14.29% | 82.93% |
| 13 | 1.88% | 4.17% | 55.00% |

When we included the SA58 nasal spray use as an independent variable in the Cox regression, the results showed that the use of SA58 nasal spray can reduce the risk of infection with HR=0.485 (95%CI: 0.354∼0.665, Figure 3)

**Figure 3.**
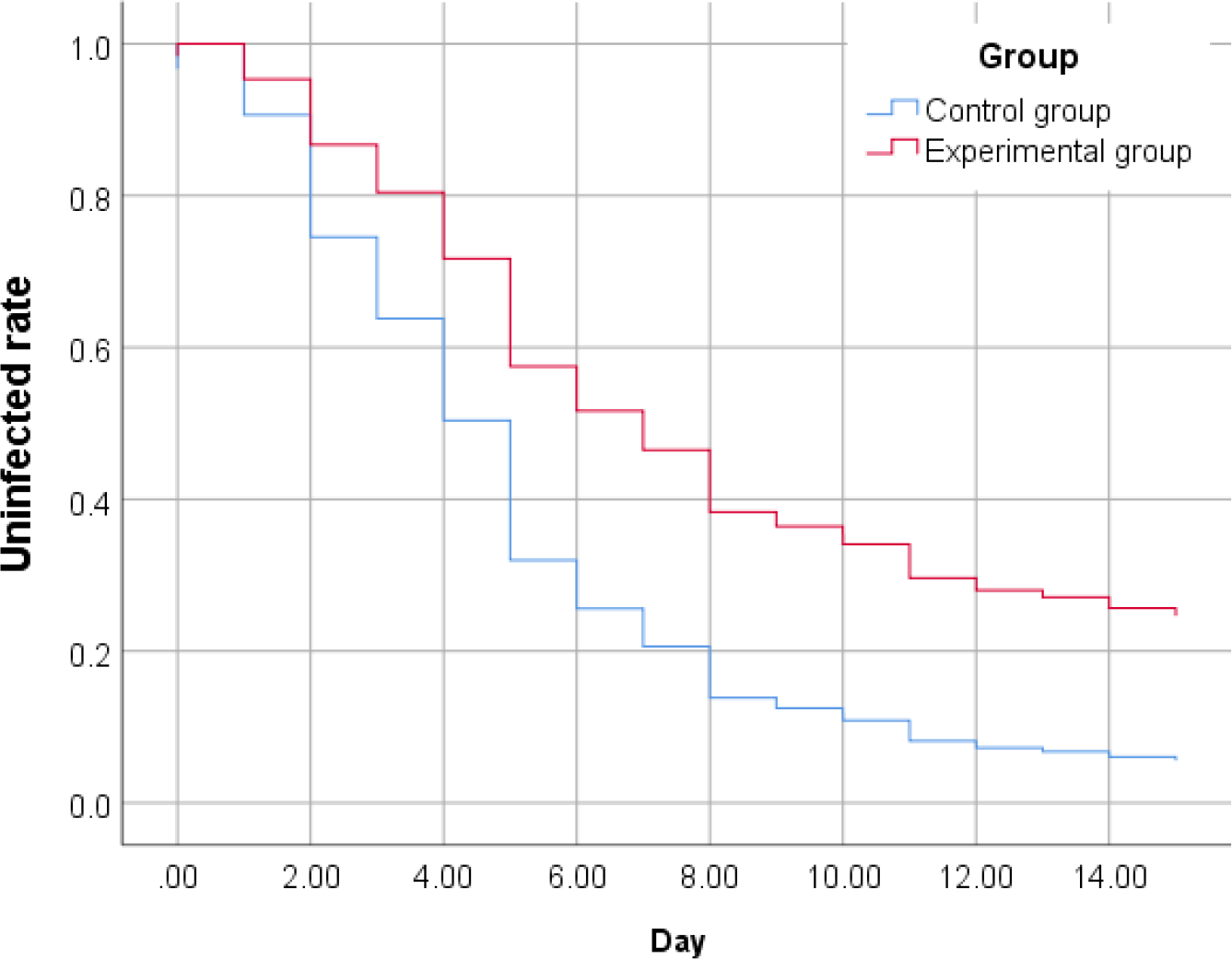
Kaplan-Meier estimates of the cumulative risk of having COVID-19.

### Safety

Of the 70 subjects who reported AEs of the SA58 nasal spray use via online questionnaire, 221 AEs were reported from 39 participants. The incidence of AEs was 55.7%. Considering some subjects may also fill in the questionnaire with COVID-19 symptoms in the early stage of infection, we separately showed the AEs occurring 2 days before positive RT-PCR results. The adjusted incidence of AEs was 41.4%. The majority of AEs included fever (25.7%), pharyngolaryngeal discomfort (24.3%), rhinorrhea (18.6%), cough (14.3%). (Figure 4). The severity of all the AEs was mild.

**Figure 4.**
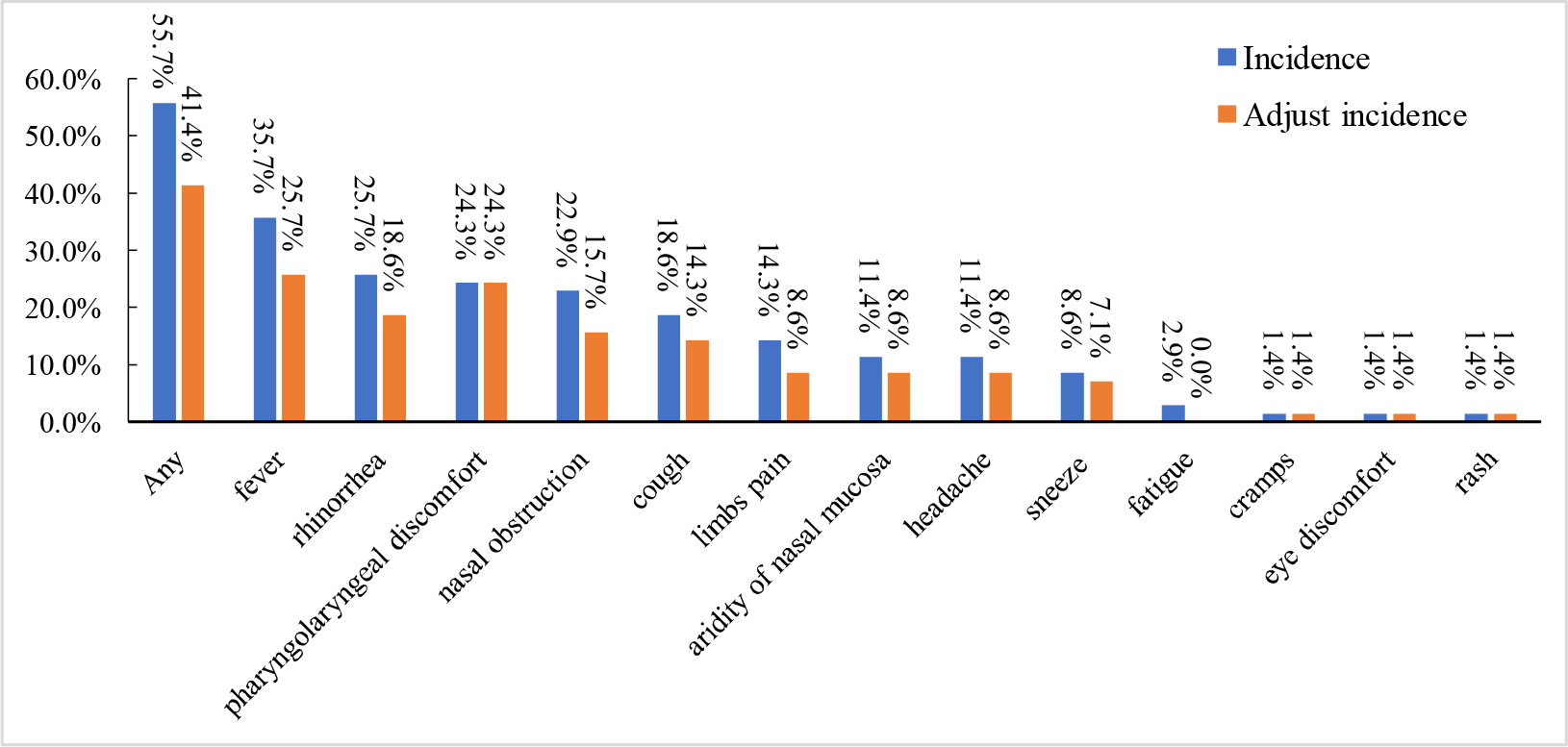
The reporting adverse events (AEs) incidence in the experimental group. Adjust incidence was calculated by excluding AEs within two days of the first positive RT-PCT which probably occurred due to COVID-19 infection.

## DISCUSSION

At the time when the study was conducted, China was adjusting its COVID-19 prevention and control policies which led to a very rapid and violent wave of SARS-CoV-2 infections mainly caused by BF.7 subvariant of Omicron. In household transmission, continuous exposure to SARS-CoV-2 usually results in a greatly increased risk of infection for family members. The observed infection rate of 95% in the control group indicates that the widespread transmission of COVID-19 is almost inevitable in a close environment with continuous exposure to SARS-CoV-2. Our preliminary results in family close contacts showed that SA58 nasal spray is moderately effective in preventing SARS-CoV-2 infection caused by Omicron BF.7 mutant with using the SA58 at least 3 times per day. Unlike the other two studies on SA58 nasal spray[12, 13], the scenario of continued exposure to COVID-19 requires people to always pay attention to use regularly the SA58 nasal spray. To minimize the influence of decreased frequency of administration on effectiveness against infection, we calculated the index called single-day effectiveness. With a frequency of 3.0 times or more per day, we found a relatively stable single-day effectiveness of 46.7%∼56.5%. The reason why single-day effectiveness was substantially higher than the total effectiveness of 33.8% was that the cumulative effectiveness can’t rule out the bias caused by decreased frequency of SA58 use. The effectiveness of SA58 nasal spray was further corroborated by the results of the Cox regression model. With the use of SA58 nasal spray as an independent variable, the hazard ratio (HR) was 0.485 (95%CI:0.354∼0.665), which means the infected risk of subjects who used the SA58 nasal spray was reduced by 51.5% (95%CI: 33.5%∼64.6%) compared to those who didn’t use SA58 at all.

The SA58 nasal spray was also shown to be well-tolerated. After we excluded the AEs occurring within 2 days before positive RT-PCR results, the incidence of AEs was 41.4%. We discovered that two of the most frequent symptoms were fever (25.7%) and pharyngolaryngeal discomfort (24.3%), which may not appear obviously linked to the use of a nasal spray. The rate of AEs was still higher than in our other two studies of SA58[12, 13], which we think is due to the inability of subjects to distinguish between adverse reactions occurring with SA58 use and COVID-19 related symptoms.

We believe that this exploratory study is of great significance. Current COVID-19 control policies of China means normal communication will return to a normal level which may not be good news for the high-risk population for COVID-19. This study demonstrated that SA58 nasal spray has a relatively good effectiveness under the scenario of continuous exposure to COVID-19, which will have a great impact on the elderly people and those with basic medical conditions at increased risk of infection. Ease of use and portability are also major benefits of the SA58 nasal spray, and it is also not burdening when used three times per day.

This study preliminarily evaluated the safety and effectiveness of SA58 nasal spray against SARS-CoV-2 family transmission and we will also conduct more in-depth and extensive studies on SA58. Considering the families recruited in this study were from Sinovac employees, their friends and relatives and almost all subjects have received one or two booster doses of COVID-19 vaccine, we didn’t evaluate the effect of previous COVID-19 vaccination on the SARS-CoV-2 infection in this study. However, the efficacy of vaccination against Omicron infection seems to be down to a very low level, as shown by the infection rate of 95% in the control group in our study. In addition, we did not assess the effect of SA58 nasal spray on COVID-19 symptoms. Considering the rapid rise of COVID-19 epidemic in China at that time and the tight medical resources, we did not finally include this indicator into this study. We will continue to investigate the efficacy of SA58 nasal spray against COVID-19 symptoms and it would be an important potential benefit for humankind to reduce the harm of COVID-19 in the future.

This study was an exploratory open-label, single-arm study. Previous studies have shown that SA58 nasal spray is highly effective in preventing COVID-19 infection, so the subjects in the experimental group were all equipped with SA58 nasal spray in order to maximize protection of subjects’ health. In addition, we conducted three telephone follow-up visits during the same period to assess how quickly the virus naturally spreads in households and try to avoid the bias caused by different periods. Our study still has a number of limitations. First, limited by the total number of RT-PCR tests per day, participants were asked to subsequently self-collect the throat swabs only every 3 days, and there is a risk of bias in the single-day effectiveness if the transmission speed is not as uniform as we assumed. Second, the time series of COVID-19 infection in the control group was obtained by subtracting the date of follow-up visits from the date of diagnosis of the first case, which may have delayed the date of infection for some subjects. This limitation would lead to slightly lower incidence in the control group early in the study, which in turn would have underestimated the early single-day effectiveness. Third, due to the high speed of this COVID-19 outbreak, a sufficient number of subjects could not be enrolled to evaluate the anti-infective effectiveness of SA58 nasal spray. In addition, subjects enrolled later may have a decline in the standardization of medication and RT-PCR as they were about to return to work.

## CONCLUSION

The use of SA58 nasal spray in family members with continuous exposure to SARS-CoV-2 has shown acceptable safety and effectiveness in avoiding SARS-CoV-2 infections. Using SA58 nasal spray three or more times a day can effectively reduce the risk of household transmission of SARS-CoV-2 by 46.7%∼56.5%.

## Data Availability

All data produced in the present study are available upon reasonable request to the authors

## Acknowledgements

We are grateful to all subjects who volunteered to participant in this trial and to all members of the clinical research teams.

